# Diagnostic serial interval as a novel indicator for contact tracing effectiveness exemplified with the SARS-CoV-2/COVID-19 outbreak in South Korea

**DOI:** 10.1101/2020.05.05.20070946

**Authors:** Sofia K. Mettler, Jihoo Kim, Marloes H. Maathuis

## Abstract

**BACKGROUND:** The clinical onset serial interval is often used as a proxy for the transmission serial interval of an infectious disease. For SARS-CoV-2/COVID-19, data on clinical onset serial intervals is limited, since symptom onset dates are not routinely recorded and do not exist in asymptomatic carriers.

**METHODS:** We define the diagnostic serial interval as the time between the diagnosis dates of the infector and infectee. Based on the DS4C project data on SARS-CoV-2/COVID-19 in South Korea, we estimate the means of the diagnostic serial interval, the clinical onset serial interval and the difference between the two. We use the balanced cluster bootstrap method to construct 95

**RESULTS:** The mean of the diagnostic serial interval was estimated to be 3.63 days (95

**CONCLUSIONS:** The relatively short diagnostic serial intervals of SARS-CoV-2/COVID-19 in South Korea are likely due to the country’s extensive efforts towards contact tracing. We suggest the mean diagnostic serial interval as a new indicator for the effectiveness of a country’s contact tracing as part of the epidemic surveillance.

## Introduction

The serial interval of an infectious disease, also known as the generation time, is defined as the time between analogous phases in successive cases of a chain of infection (Porta, 2016). The *transmission interval*, or the time between the infection events of the infector and the infectee, is a particularly important serial interval as it determines how rapidly the disease can spread in the community (Fine, 2003) and provides a time window for its containment. This interval, however, is difficult to measure, as times of infection are often unknown. A commonly used alternative is the *clinical onset serial interval*, or the time between the onset of symptoms of the infector and the infectee (Fine, 2003). The clinical onset serial interval is easier to measure than the transmission interval, as the time of symptom onset is more frequently known than the time of infection.

There is a rapidly growing literature on the clinical onset serial interval of COVID-19, with reported mean values ranging from 3.96 to 7.5 days. Notable examples are an early study from Wuhan, China by Li et al. (2020), reporting a mean of 7.5 days based on 6 infector-infectee pairs, an early meta-analysis by Nishiura, Linton, et al. (2020), reporting a median of 4.0 days based on 28 pairs, as well as the first larger data analysis by Du et al. (2020), reporting a mean of 3.96 days based on 468 pairs. Two South Korean studies on the clinical onset serial interval of COVID-19 are currently available, with Ki (2020) reporting a mean of 6.6 days based on 9 pairs, and Son et al. (2020) estimating a mean of 5.54 days based on 28 pairs.

The clinical onset serial interval has some limitations. First, it is undefined if the infector or the infectee remains asymptomatic. Second, the clinical onset serial interval relies on subjective perception and accurate reporting of the onset of symptoms by patients. Finally, the time of symptom onset is not always documented.

We therefore introduce a new type of serial interval, the *diagnostic serial interval*, which we define as the time between the diagnosis dates of the infector and the infectee. In this study of SARS-CoV-2/COVID-19, we define the diagnosis to be laboratory confirmation of SARS-CoV-2 infection, regardless of the onset of symptoms.

The diagnostic serial interval is easier to determine than the clinical onset serial interval, since dates of diagnosis are routinely recorded and less subjective than the onset of symptoms. Moreover, in contrast to the clinical onset serial interval, it is defined for asymptomatic carriers of SARS-CoV-2. This is especially favorable as asymptomatic transmission appears likely (Rothe et al., 2020), and a significant portion of asymptomatic carriers remain asymptomatic (Nishiura, Kobayashi, et al., 2020; Zhou et al., 2020).

While the clinical onset serial interval is largely a characteristic of the pathogen, we argue that the diagnostic serial interval contains additional information on public health policy and capacity. In particular, a well-functioning contact tracing system leads to shorter diagnostic serial intervals, which can in turn contribute to breaking chains of infections (see Figure 1). We therefore propose to use the mean diagnostic serial interval as a novel indicator for the effectiveness of contact tracing, besides other indicators like the proportion of cases with unknown transmission routes.

**Figure 1:**
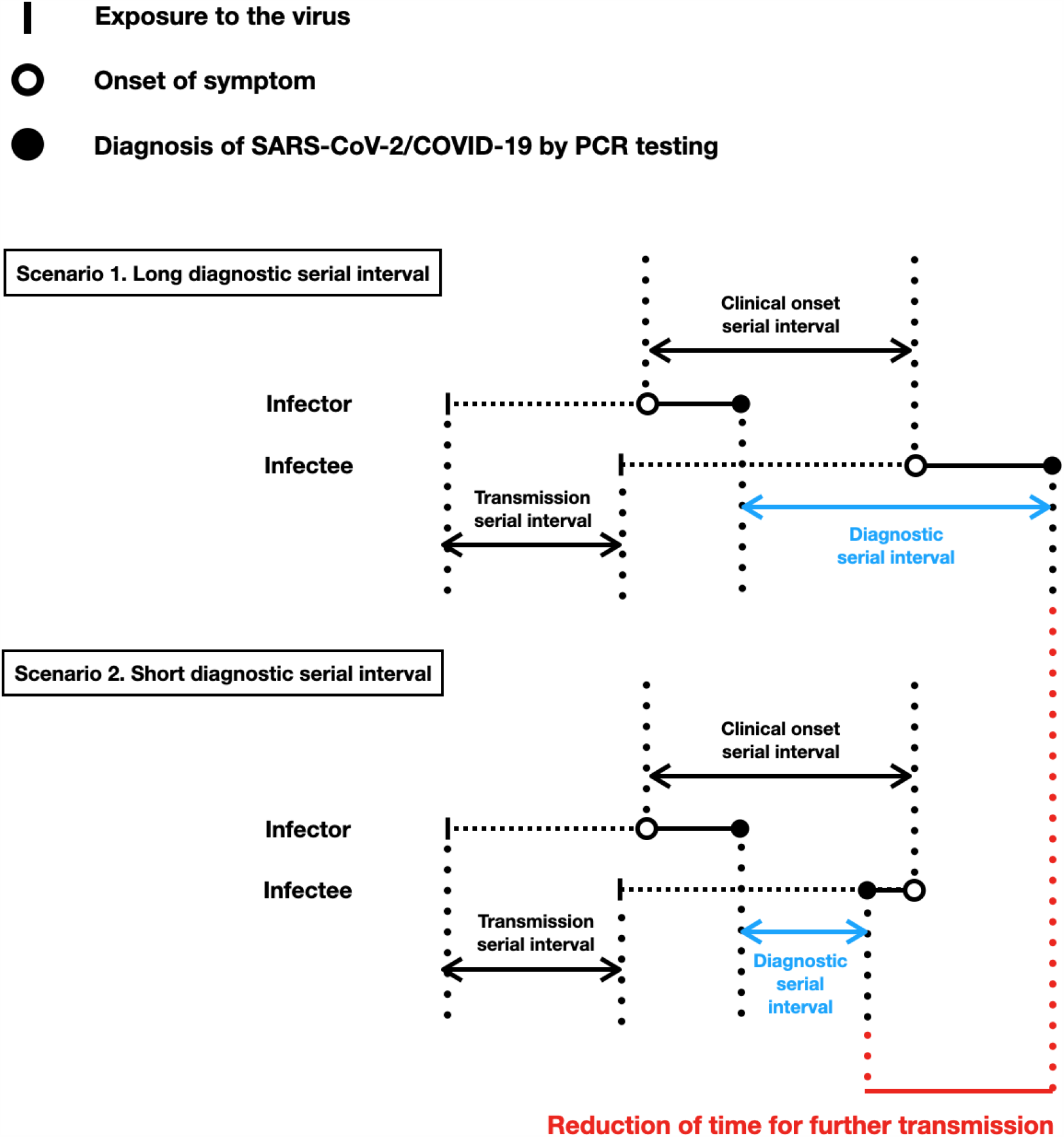
Graphical illustration of the diagnostic serial interval and the potential of short diagnostic serial intervals to reduce further transmission.

We study the clinical onset serial interval and the newly introduced diagnostic serial interval of SARS-CoV-2/COVID-19, using a rich data set of infector-infectee pairs observed in South Korea between January 20th and June 30th, 2020.

## Data and Methods

### Data and inclusion criteria

A total of 12,850 individuals were confirmed to be infected with SARS-CoV-2 in South Korea between January 20th and June 30th (Korea Centers for Disease Control & Prevention, 2020). The seventeen regional governments of South Korea have published daily information on newly infected individuals, including their date of diagnosis, age, gender, infection route, infector (if known) and symptom onset date (if symptomatic and reported by the time of diagnosis). Among the seventeen regional governments, sixteen have published information on over 90% of their confirmed cases, the only exception being Daegu that published information on only 2% of its 6,906 patients until June 30th (Korea Centers for Disease Control & Prevention, 2020). Our data set was obtained in collaboration with the DS4C project (Kim et al., 2020). It contains information on 5,201 individuals who were confirmed to be infected with SARS-CoV-2 between January 20th and June 30th, 2020. Figure 2 shows the daily new cases of SARS-CoV-2/COVID-19 in South Korea and the data coverage by date. The data coverage by region is shown in Supplement A. The data used in this analysis are available from our data depository (h<u>ttps://github.com/DSI-COVID/DS4C0701.git).</u>

**Figure 2:**
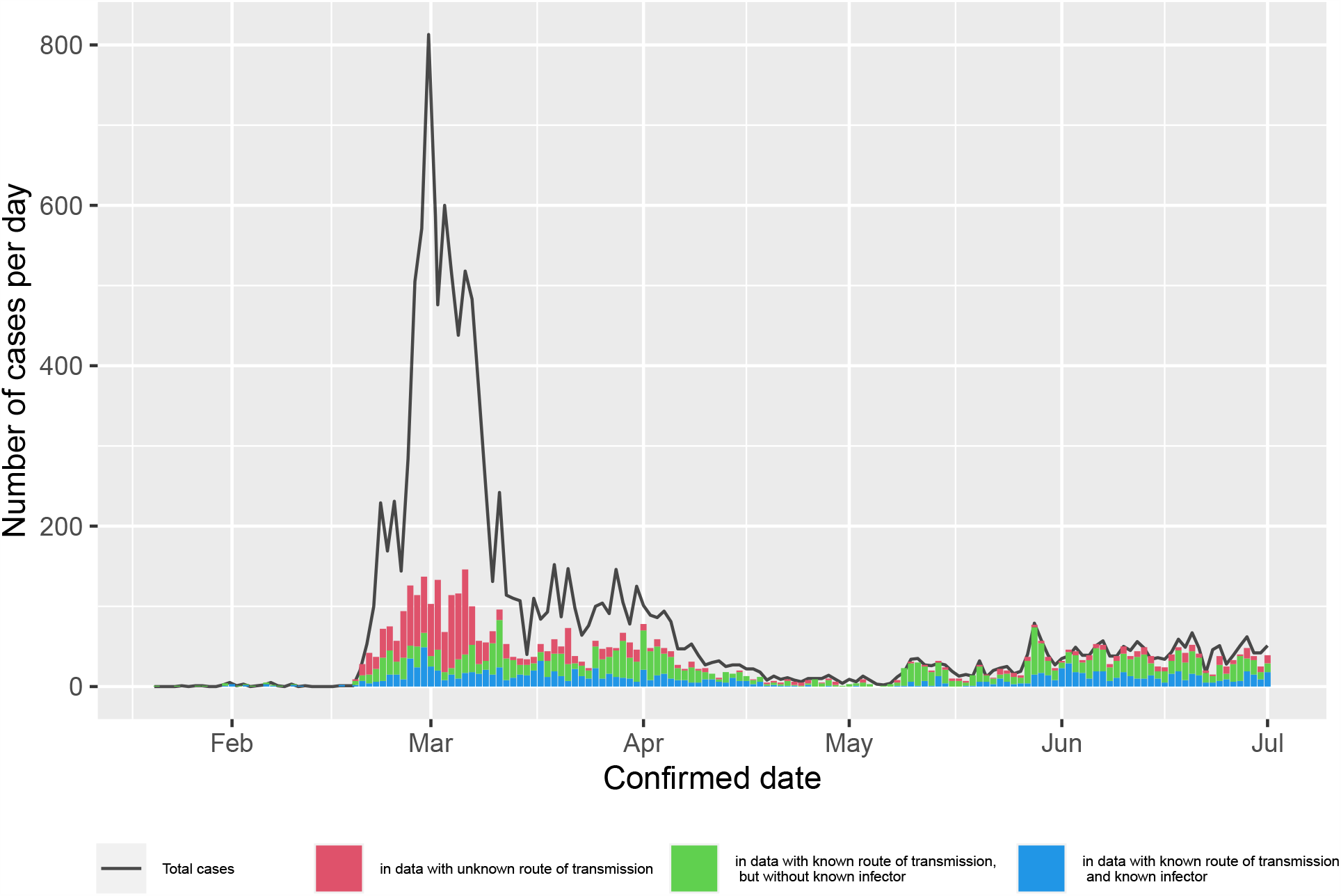
Daily new cases of SARS-CoV-2/COVID-19 in South Korea between January 20th and June 30th, 2020.

Among these 5,201 individuals, there were 1,360 known infector-infectee pairs. The date of diagnosis was recorded for all infectors and infectees in these pairs. In order to reduce bias resulting from right truncation, we excluded 125 pairs for which the infector’s diagnosis date was later than June 16th. The remaining 1,235 infector-infectee pairs, containing 555 unique infectors and 1,235 unique infectees, were used to study the diagnostic serial interval. We refer to these data as data set A.

Twelve of the seventeen regional governments have sporadically reported symptom-related information including the symptom onset dates. Among the 1,360 known infector-infectee pairs, there were 106 pairs for which the onset date of symptoms was known for both the infector and the infectee. Again, in order to reduce bias from right truncation, we excluded 4 pairs for which the infector’s symptom onset date was later than June 16th. The remaining 102 infector-infectee pairs, containing 60 unique infectors and 102 unique infectees, were used to study the diagnostic serial interval, the clinical onset serial interval and the difference between the two. We refer to these data as data set B.

The data preparation is described in Supplement B, together with the corresponding R code.

### Statistical Methods

Based on data set A (1,235 infector-infectee pairs) we estimated the mean of the diagnostic serial interval. Based on data set B (102 infector-infectee pairs with known symptom onset dates) we estimated the mean of the diagnostic serial interval, the clinical onset serial interval and the difference between the two (diagnostic serial interval*−*clinical onset serial interval). The latter was done using a matched sample (paired) analysis. We computed 95% confidence intervals for these parameters using the bootstrap method. To account for dependencies between observations caused by common infectors, we used the balanced cluster bootstrap method which regards each infector as a cluster (Davison et al., 1997).

We also considered robust estimation of all parameters using trimmed means with different degrees of trimming (2.5%, 5% and 12.5%) as well as different types of bootstrap confidence intervals (normal-based, reversed percentile and percentile).

All computations were performed using the statistical software R. All R code is provided in Supplement D.

## Results

Figure 3 visualizes the relationship between infectors’ and infectees’ symptom onset dates (Figure 3(a)) and diagnosis dates (Figure 3(b)). The data points above the diagonal line in Figure 3(a) show that some infectees developed symptoms before their infectors, indicating the presence of asymptomatic transmission. A similar phenomenon occurs for the diagnosis dates in Figure 3(b), but to a lesser extent.

**Figure 3:**
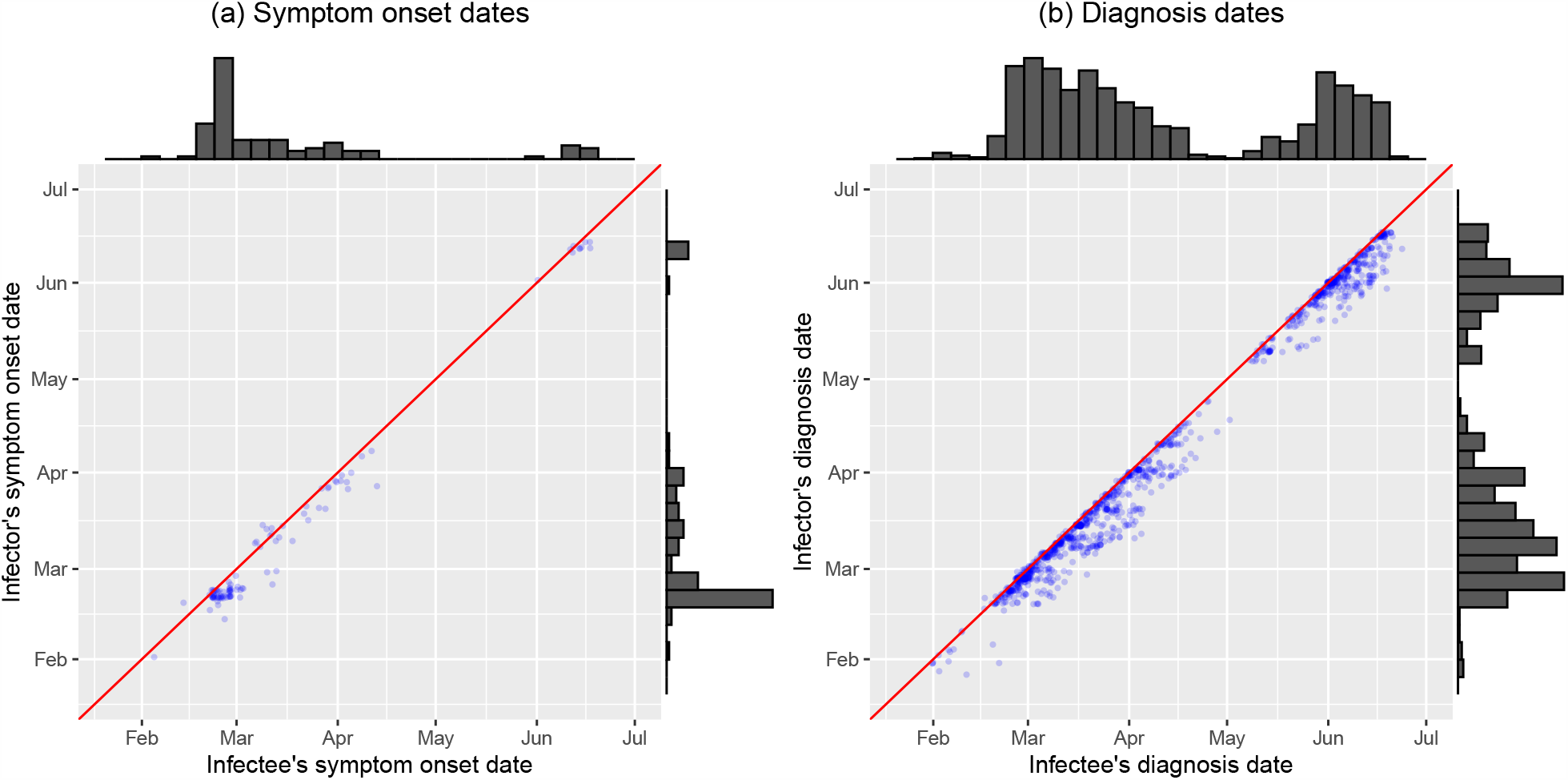
(a) Scatterplot of the symptom onset dates of infector-infectee pairs. Each point corresponds to an infector-infectee pair. Histograms of the symptom onset dates of infectees and infectors are displayed at the horizontal and vertical margin, respectively. (b) Scatterplot of the diagnosis dates of infector-infectee pairs, analogous to (a).

Figure 4 show histograms of the diagnostic serial interval, the clinical onset serial interval and their difference. Table 1 show the corresponding estimated means with 95% normal-based bootstrap confidence intervals.

**Figure 4:**
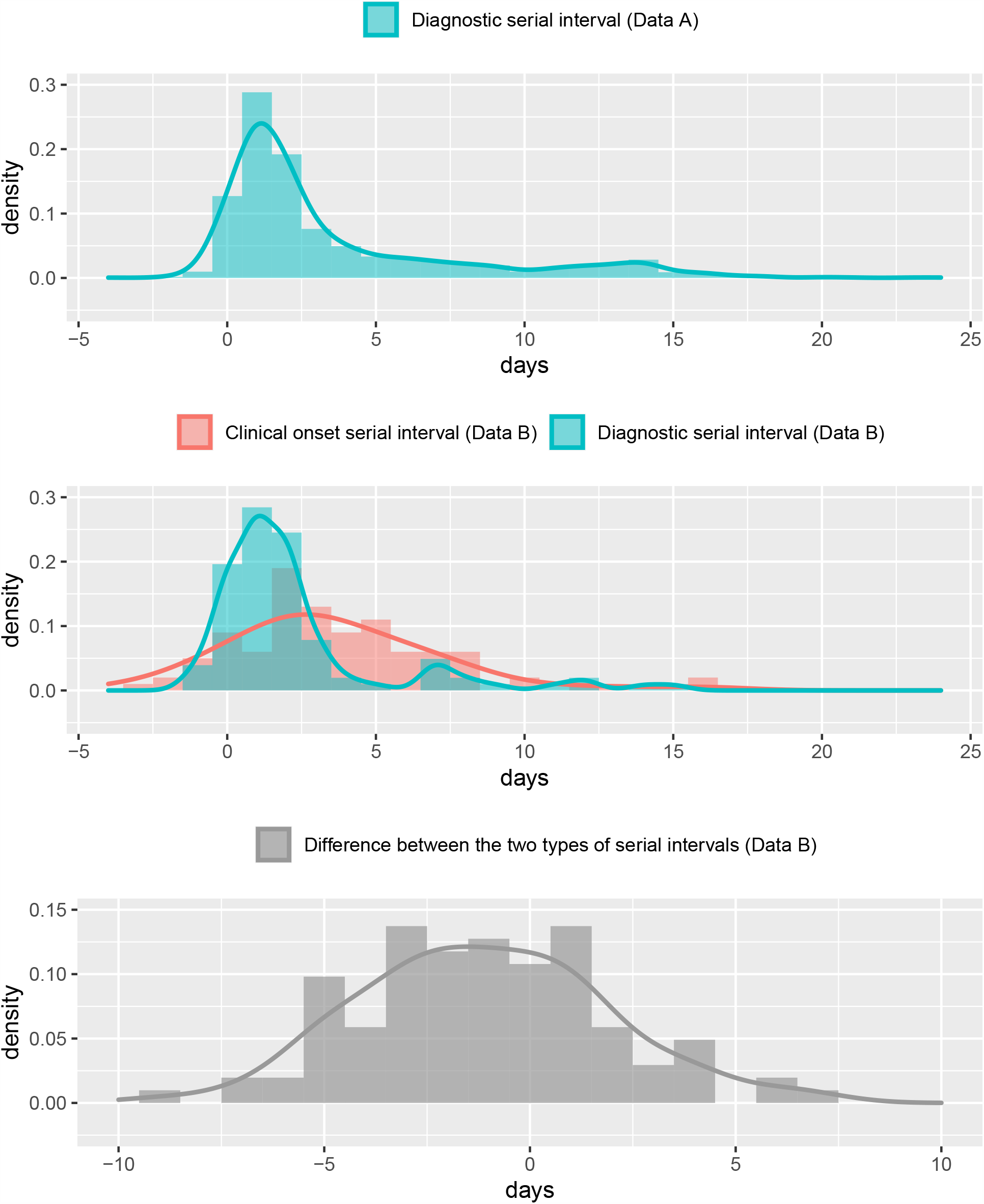
(a) Histogram of the diagnostic serial interval (data set A, blue) in days. (b) Histograms of the clinical onset serial interval (data set B, red) and the diagnostic serial interval (data set B, blue) in days. (c) Histogram of the difference (diagnostic serial interval - clinical onset serial interval) (data set B, gray). In all plots, lines show the estimated density functions.

We first discuss the results for data set A. The histogram of the diagnostic serial interval (Figure 4(a)) shows that the distribution has a heavy right tail. Among the 1,235 pairs in this data set, there were 15 pairs (1.21

%) in which infectees were diagnosed before their infectors, resulting in a negative diagnostic serial interval. The estimated mean diagnostic serial interval was 3.63 days (95% CI: 3.24, 4.01), which is relatively short in comparison with the previously reported mean or median clinical onset serial intervals. The width of the confidence interval is short due to the relatively large data set.

We now discuss the results for data set B. The histograms in Fig 4(b) show that the distributions of the clinical onset serial interval and the diagnostic serial interval appear to have different shapes. Among the 102 pairs in this data set, there were 11 pairs (10.78 %) with a negative clinical onset serial interval and 4 pairs (3.92 %) with a negative diagnostic serial interval. The mean clinical onset serial interval was estimated to be 3.43 days (95% CI: 2.62, 4.24), which is rather short compared to the previously reported mean values of the clinical onset serial interval. The mean diagnostic serial interval based on data set B was estimated to be 2.31 days (95% CI: 1.48, 3.14). This is considerably shorter than the mean diagnostic serial interval based on data set A, indicating that data set B is a special subset of data set A.

A histogram of the difference between the two intervals (diagnostic serial interval - clinical onset serial interval) for data set B is shown in Figure 4(c). Among the 102 pairs in this data set, 60 pairs (58.82%) had a shorter diagnostic serial interval than clinical onset serial interval. The matched sample analysis yielded an estimated mean difference of -1.12 days (95% CI: -1.98, −0.26). This difference is significant at the *α* = 0.05 level, indicating that the diagnostic serial intervals are significantly shorter than their corresponding clinical onset serial intervals.

Results for trimmed means and different types of bootstrap confidence intervals are shown in Supplement C. As expected due to the long right tails of the distributions, the trimmed means become smaller when a larger proportion of the data is trimmed. The different types of bootstrap confidence intervals do not make a large difference.

## Discussion

We introduced the diagnostic serial interval, that is, the time between the dates of diagnosis of the successive cases in a chain of infection. The diagnostic serial interval is especially relevant for SARS-CoV-2/COVID-19, as asymptomatic carriers appear to be able to infect others before the onset of symptoms. Therefore, preventive measures have to be taken before symptoms develop in order to be able to break the infection chain. While the clinical onset serial interval is based on symptoms, the diagnostic serial interval is based on the detectablility of the antigen, i.e., viral load, which is likely to be more relevant for viral shedding and transmission. Short diagnostic serial intervals imply faster identification and isolation of new infection events and therefore less time for further transmission (see Figure 1).

We analyzed a rich data set of SARS-CoV-2/COVID-19 cases in South Korea. The mean of the diagnostic serial interval was estimated to be 3.63 days (95% CI: 3.24, 4.01). The diagnostic serial interval was shown to be significantly shorter than the clinical onset serial interval (estimated mean difference -1.12 days, 95% CI: -1.98, −0.26). The relatively short diagnostic serial interval in South Korea is likely due to the country’s intensive efforts in contact tracing.

Our analysis has several limitations, the main one being that our data do not constitute a random sample of the population of infected individuals. As a result, we may suffer from selection bias in various ways. For example, the province of Daegue, which was severely affected by the epidemic, is strongly underrepresented. Also, some selection bias for shorter intervals is likely to exist, as infector-infectee pairs with shorter serial intervals may be identified more easily than those with longer serial intervals. Another limitation of our study is the potential dependency among observations. While the cluster bootstrap method accounts for dependencies between pairs with a common infector, it does not account for dependencies caused by long chains of infection or different viral subtypes with varying virulence and infectivity.

In countries that managed to contain early outbreaks, concerns for a second wave are rising. Although the most powerful containment measures such as a lockdown, international travel bans and strict social distancing appear to have contributed to the containment of the earlier outbreaks, these measures are not sustainable in the long run. Contact tracing is an important tool that is sustainable, but its effectiveness cannot be merely assessed by the size of the epidemic, its growth rate or the mortality, as many other factors play a role in the dynamic of the epidemic. We therefore suggest that the diagnostic serial interval, in addition to other measures such as the proportion of cases with unknown transmission routes, can serve as a novel indicator for the effectiveness of contact tracing. This indicator could be compared over time or among regions.

It is noteworthy that infector-infectee relations are rather infrequently known in our data set, even among cases with known transmission routes. This is mainly due to cluster infection events in which multiple individuals may act as infectors or infectees. In such cases, the transmission interval, the clinical onset serial interval and the diagnostic serial interval cannot be calculated. We may consider a cluster-specific definition of the diagnostic serial interval by assuming the first confirmed case in the cluster to be the infector of all others in the cluster. Although this definition may lead to an overestimation of the true diagnostic serial interval, it may nevertheless be useful in assessing the effectiveness of contact tracing.

**Table 1:**
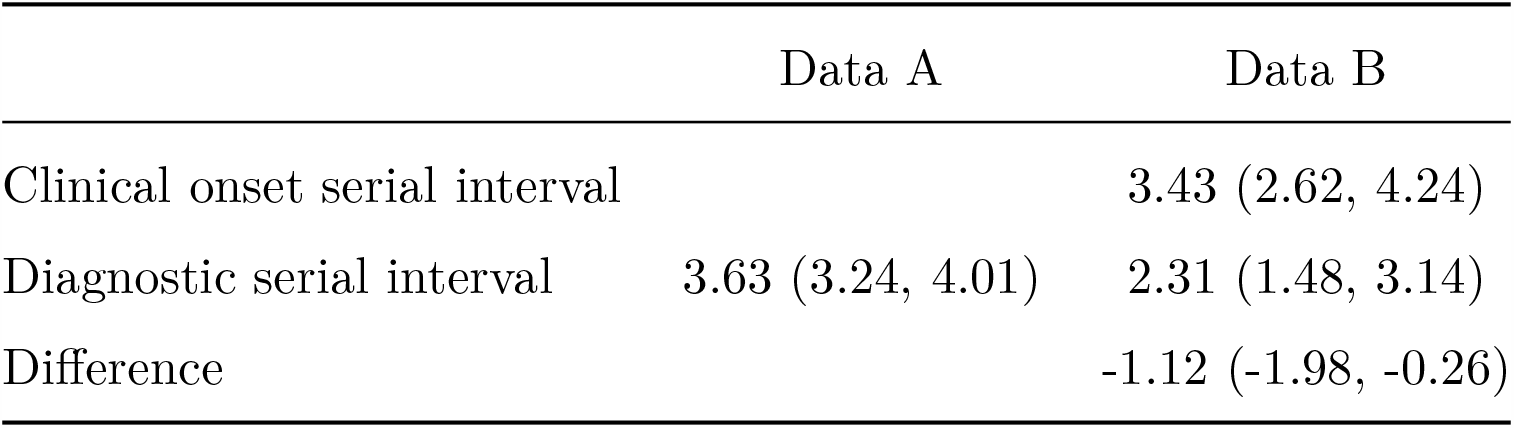
Estimated means of the clinical onset serial interval, the diagnostic serial interval, and their difference (diagnostic serial interval - clinical onset serial interval) of SARS-CoV-2/COVID-19 in South Korea, including 95% normal-based bootstrap confidence intervals (in days).

## Data Availability

Our research is based on the publicly accessible data which are part of the DS4C project. The data set used in the study will be made available once the article is peer-reviewed and published.

## Appendix

## Acknowledgements

We thank Prof. Jan Fehr at the Department of Public & Global Health at the University of Zurich and Prof. Werner Stahel at the Seminar for Statistics at ETH Zurich for helpful discussions.

## Footnote

Sofia Mettler is a candidate of medicine at the Faculty of Medicine at the University of Zurich and a master’s candidate in Statistics at ETH Zurich. Jihoo Kim is the chief director of the DS4C project and a master’s candidate in Computer Science at Hanyang University. Prof. Marloes Maathuis is Professor of Statistics at ETH Zurich.

### Patient or Public Involvement Statement and Ethics Committee Approval

Our research is based on publicly available data and does not require to involve patients or the public in the design, or conduct, or reporting, or dissemination plans. Our research does not require ethics committee approval.

### Conflict of interest statement

All authors have completed the ICMJE uniform disclosure form and declare: no support from any organization for the submitted work; no financial relationships with any organizations that might have an interest in the submitted work in the previous three years, no other relationships or activities that could appear to have influenced the submitted work.

### Author statement

Sofia Mettler conceived the topic, conducted literature research, and wrote the manuscript and R code. Jihoo Kim collected and tailored the data for our analysis. Prof. Marloes Maathuis suggested statistical methodology, wrote the manuscript and supervised the entire work.

## Supplementary Material

### Supplement A. The size of SARS-CoV-2/COVID-19 outbreak and data coverage by region

**Figure 5:**
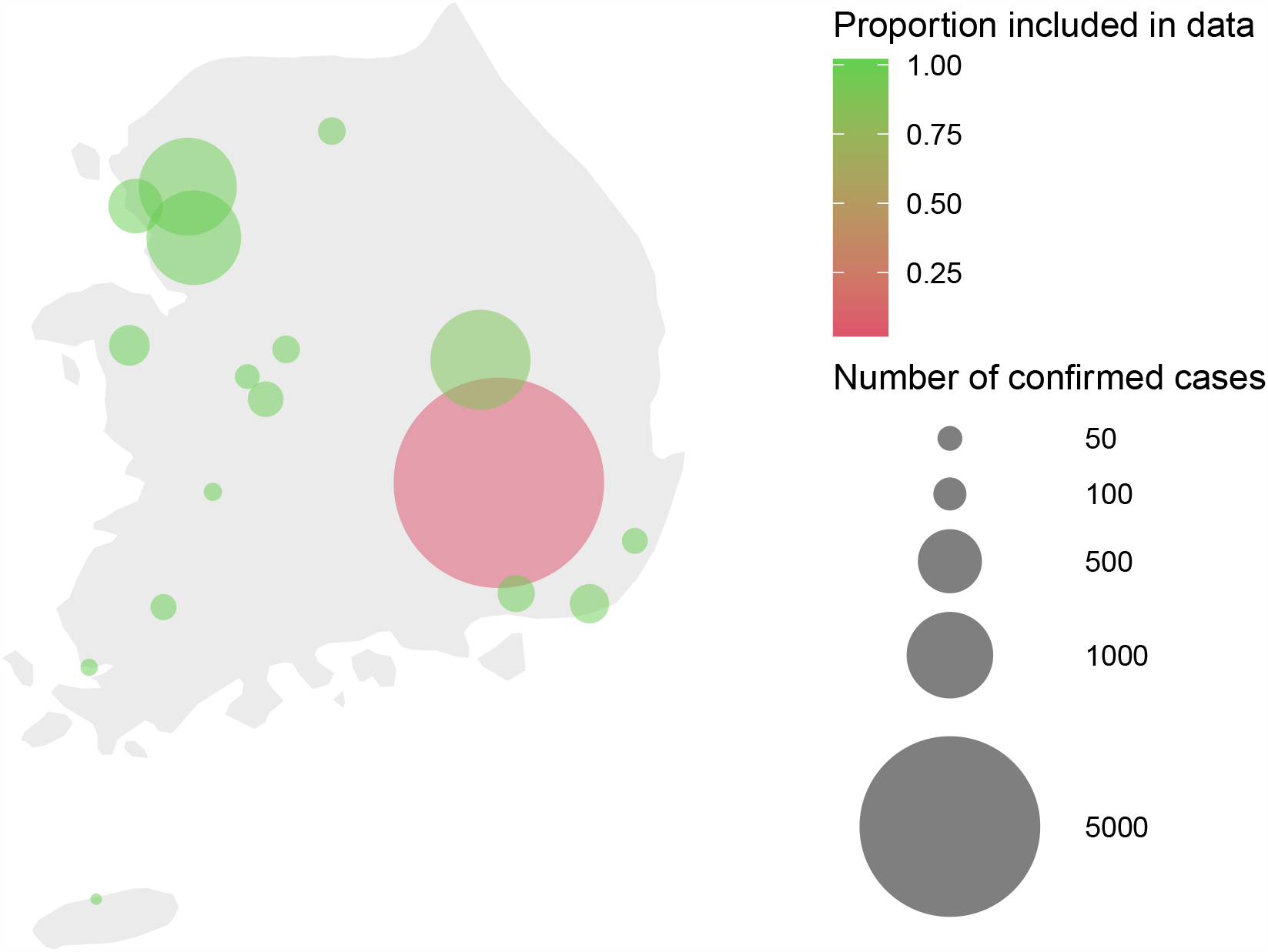
Map of South Korea indicating the size of the SARS-CoV-2/COVID-19 outbreak and the data coverage by region as of June 30th, 2020

**Table 2:** The size of the SARS-CoV-2/COVID-19 outbreak and data coverage by region in South Korea as of June 30th. The update of the total number of cases by region may be slower than the update of the information on confirmed cases, leading to a larger number of cases included in our data than the total number in this table.

| Province | Total cases | In data | Data coverage |
| --- | --- | --- | --- |
| Busan | 154 | 151 | 0.981 |
| Chungcheongbuk-do | 65 | 64 | 0.985 |
| Chungcheongnam-do | 168 | 169 | 1.006 |
| Daegu | 6907 | 137 | 0.020 |
| Daejeon | 121 | 119 | 0.983 |
| Gangwon-do | 65 | 63 | 0.969 |
| Gwangju | 56 | 56 | 1.000 |
| Gyeonggi-do | 1223 | 1225 | 1.002 |
| Gyeongsangbuk-do | 1389 | 1252 | 0.901 |
| Gyeongsangnam-do | 134 | 133 | 0.993 |
| Incheon | 343 | 343 | 1.000 |
| Jeju-do | 19 | 19 | 1.000 |
| Jeollabuk-do | 27 | 27 | 1.000 |
| Jeollanam-do | 25 | 25 | 1.000 |
| Sejong | 50 | 51 | 1.020 |
| Seoul | 1321 | 1312 | 0.993 |
| Ulsan | 55 | 55 | 1.000 |

### Supplement B. Data preparation

**Inclusion criteria for analysis**

**Figure 6:**
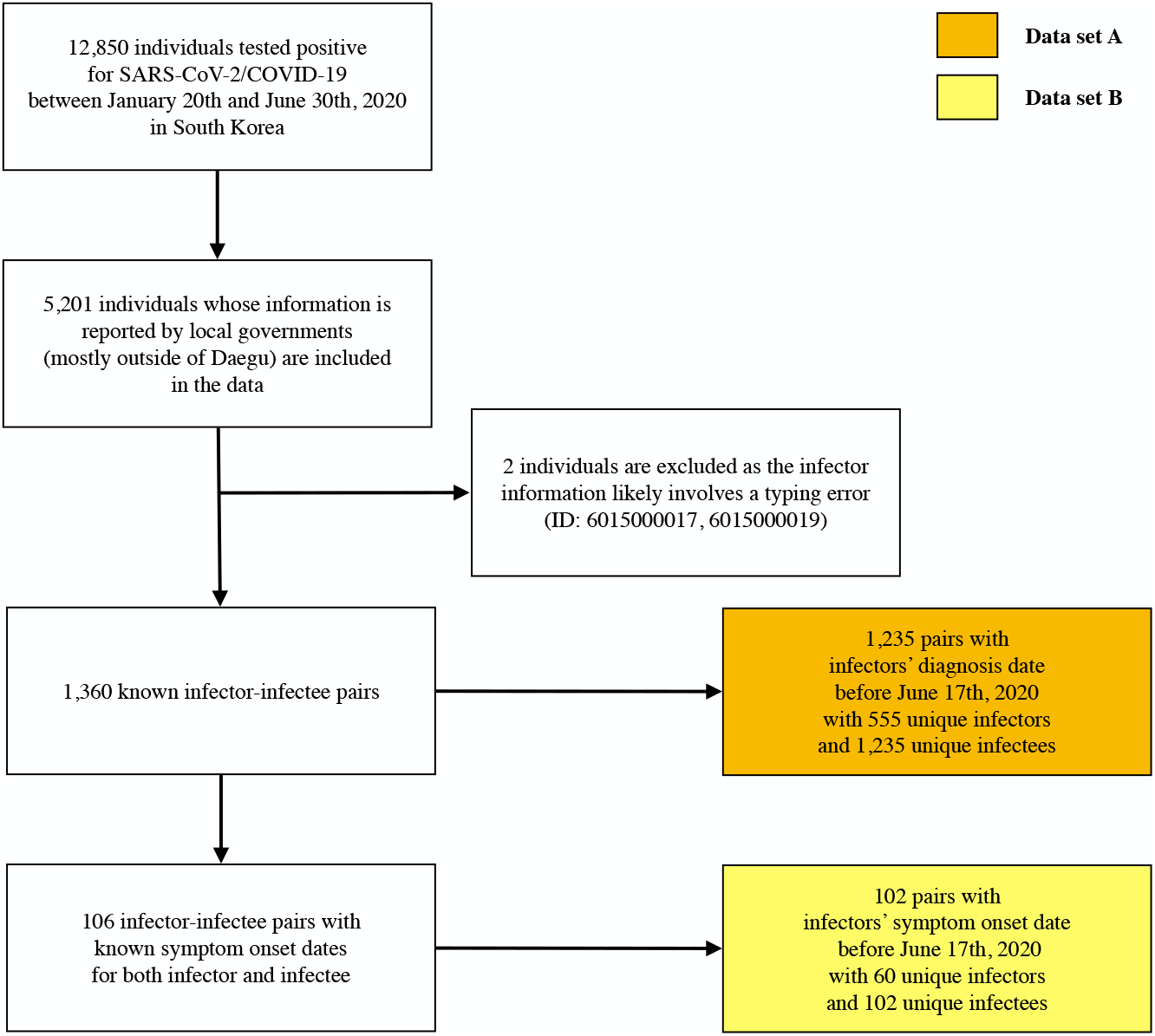
Inclusion criteria for analysis

**Data preparation code**

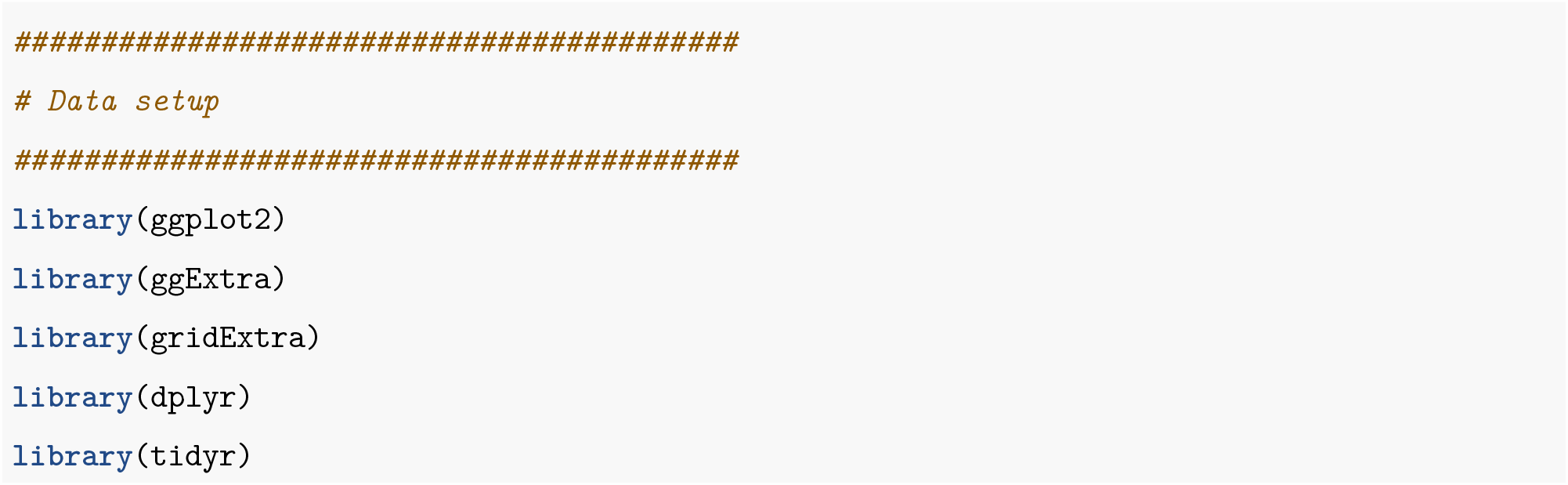
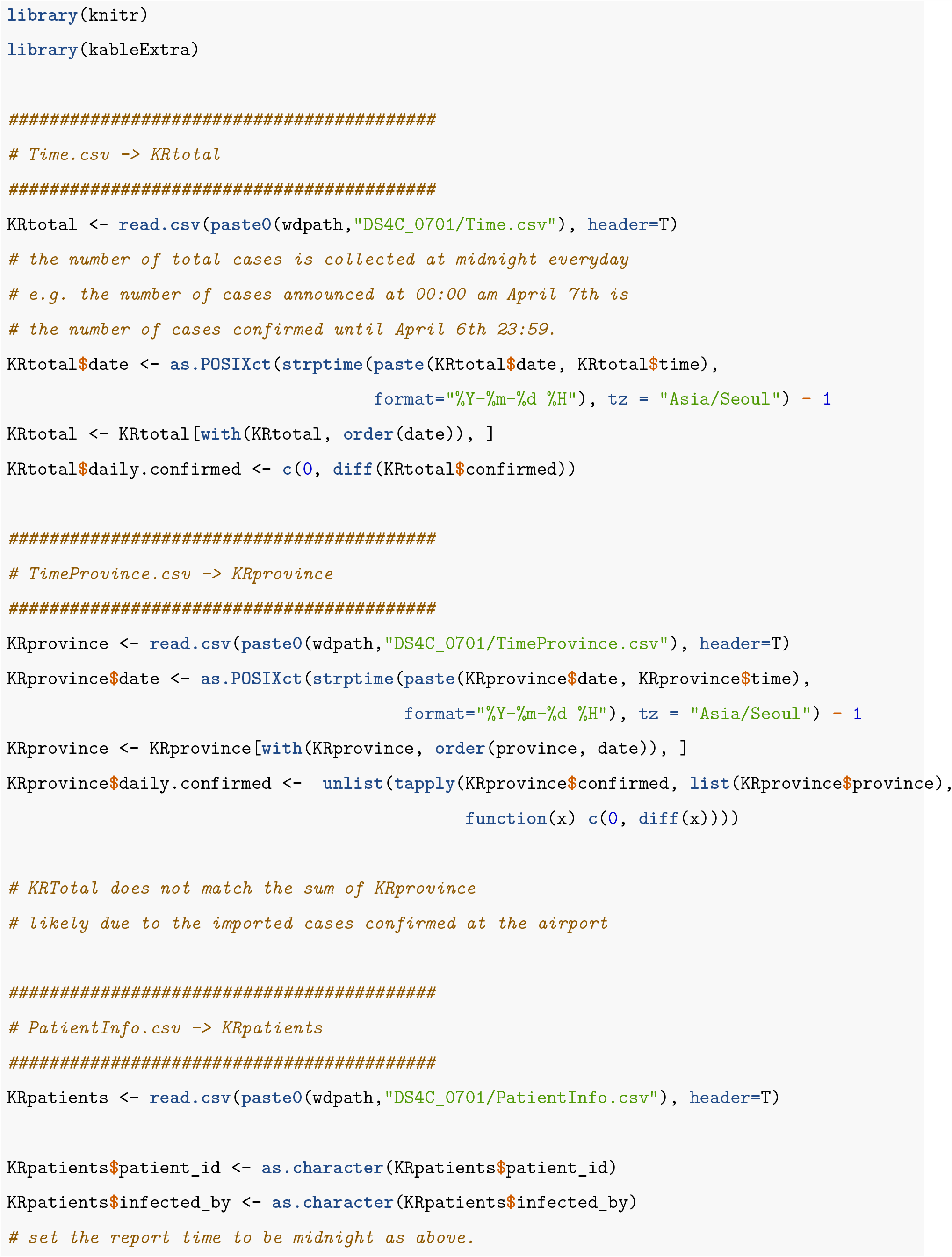
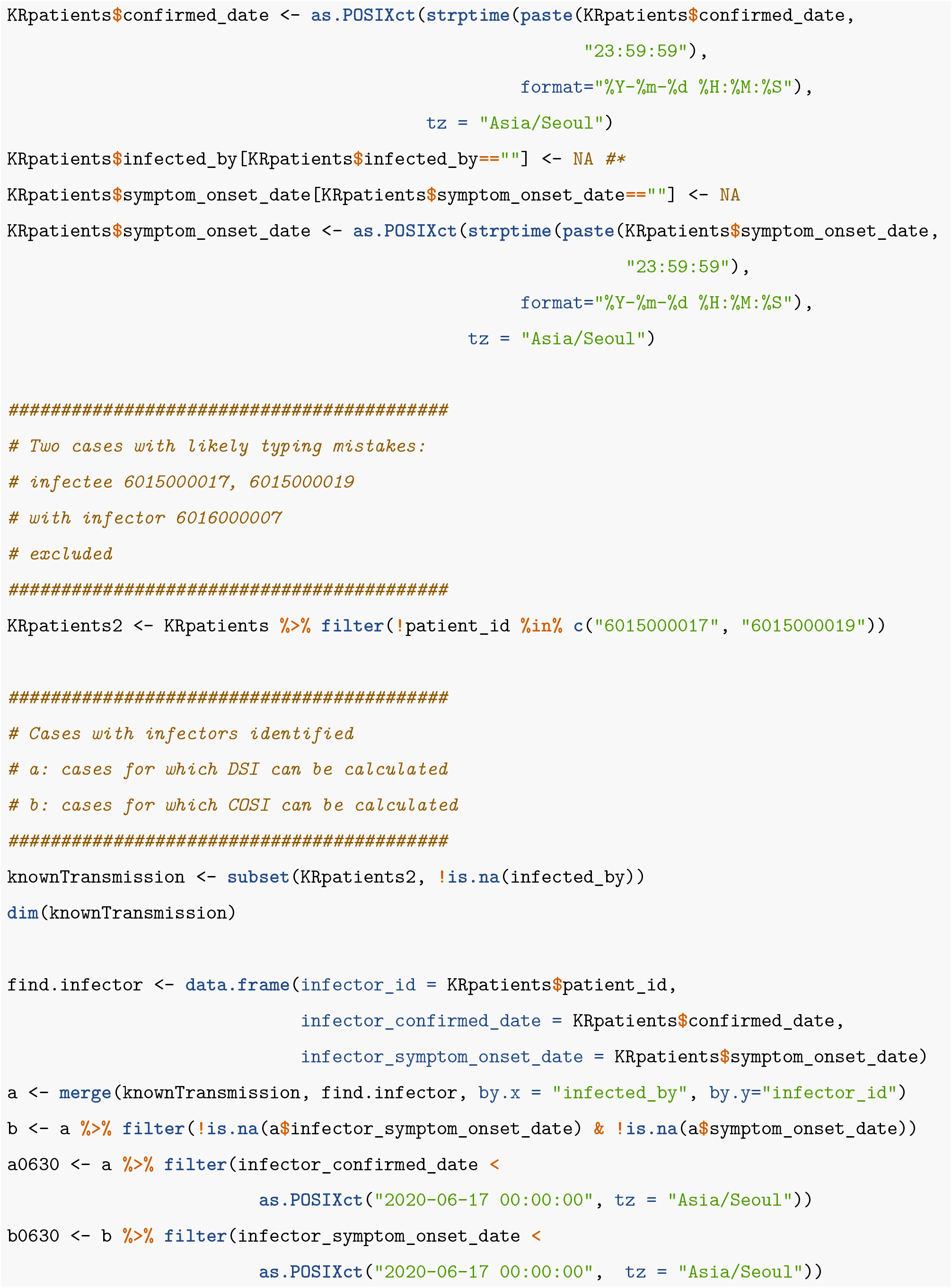

### Supplement C. Bootstrap results using trimmed means and different types of bootstrap confidence intervals

Bootstrap results using trimmed means are shown below. The first column *trim* in each table indicates the proportion of observations that are trimmed at each end. For example, the value of 0.025 indicates that 2.5% of observations at each end, i.e. 5% of observations in total, are excluded when computing the mean. The lower and upper boundaries of the 95% normal-based, reversed percentile and percentile bootstrap confidence intervals are also shown in each table.

**Table 3:** Diagnostic Serial interval (Data A)

| trim | mean | Lower.normal | Upper.normal | Lower.reversepercentile | Upper.reversepercentile | Lower.percentile | Upper.percentile |
| --- | --- | --- | --- | --- | --- | --- | --- |
| 0.000 | 3.63 | 3.24 | 4.01 | 3.23 | 4.00 | 3.25 | 4.02 |
| 0.025 | 3.39 | 3.00 | 3.78 | 2.99 | 3.77 | 3.01 | 3.79 |
| 0.050 | 3.19 | 2.78 | 3.59 | 2.77 | 3.58 | 2.79 | 3.60 |
| 0.125 | 2.63 | 2.23 | 3.03 | 2.20 | 3.00 | 2.25 | 3.05 |

**Table 4:** Clinical Onset Serial interval (Data B)

| trim | mean | Lower.normal | Upper.normal | Lower.reversepercentile | Upper.reversepercentile | Lower.percentile | Upper.percentile |
| --- | --- | --- | --- | --- | --- | --- | --- |
| 0.000 | 3.43 | 2.62 | 4.24 | 2.65 | 4.29 | 2.57 | 4.21 |
| 0.025 | 3.36 | 2.55 | 4.17 | 2.56 | 4.21 | 2.51 | 4.15 |
| 0.050 | 3.29 | 2.51 | 4.08 | 2.54 | 4.12 | 2.47 | 4.05 |
| 0.125 | 3.23 | 2.46 | 4.00 | 2.55 | 4.07 | 2.40 | 3.91 |

**Table 5:** Diagnostic Serial interval (Data B)

| trim | mean | Lower.normal | Upper.normal | Lower.reversepercentile | Upper.reversepercentile | Lower.percentile | Upper.percentile |
| --- | --- | --- | --- | --- | --- | --- | --- |
| 0.000 | 2.31 | 1.483 | 3.14 | 1.302 | 2.96 | 1.66 | 3.33 |
| 0.025 | 2.13 | 1.288 | 2.98 | 1.076 | 2.75 | 1.51 | 3.19 |
| 0.050 | 1.91 | 1.111 | 2.71 | 0.850 | 2.43 | 1.40 | 2.98 |
| 0.125 | 1.58 | 0.928 | 2.23 | 0.686 | 1.98 | 1.17 | 2.47 |

**Table 6:** Difference between clinical onset and diagnostic serial intervals (Data B)

| trim | mean | Lower.normal | Upper.normal | Lower.reversepercentile | Upper.reversepercentile | Lower.percentile | Upper.percentile |
| --- | --- | --- | --- | --- | --- | --- | --- |
| 0.000 | -1.12 | -1.98 | -0.259 | -2.08 | -0.397 | -1.84 | -0.158 |
| 0.025 | -1.13 | -2.02 | -0.250 | -2.12 | -0.392 | -1.87 | -0.147 |
| 0.050 | -1.15 | -2.06 | -0.246 | -2.16 | -0.391 | -1.91 | -0.149 |
| 0.125 | -1.21 | -2.16 | -0.254 | -2.25 | -0.420 | -1.99 | -0.156 |

### Supplement D. R code

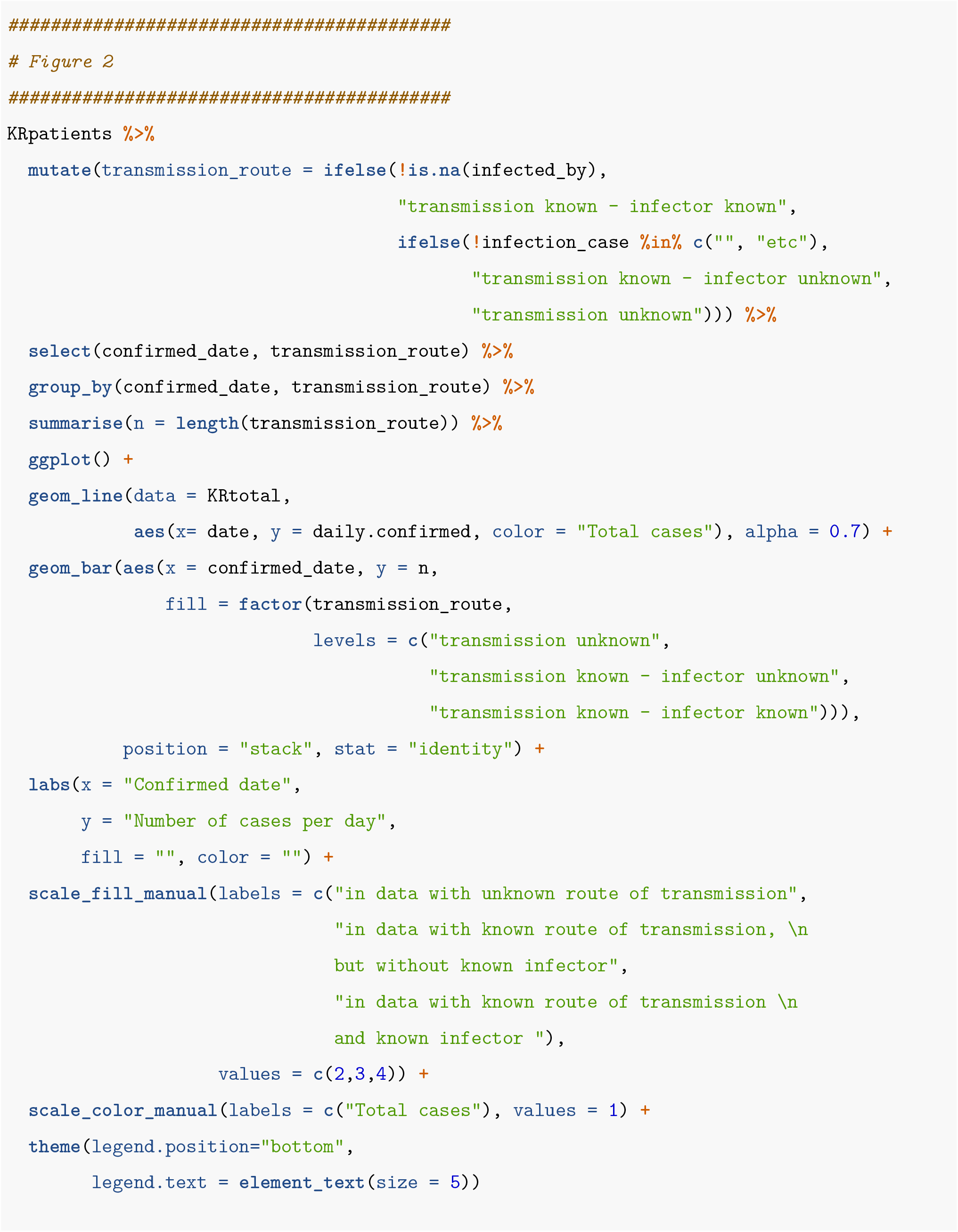
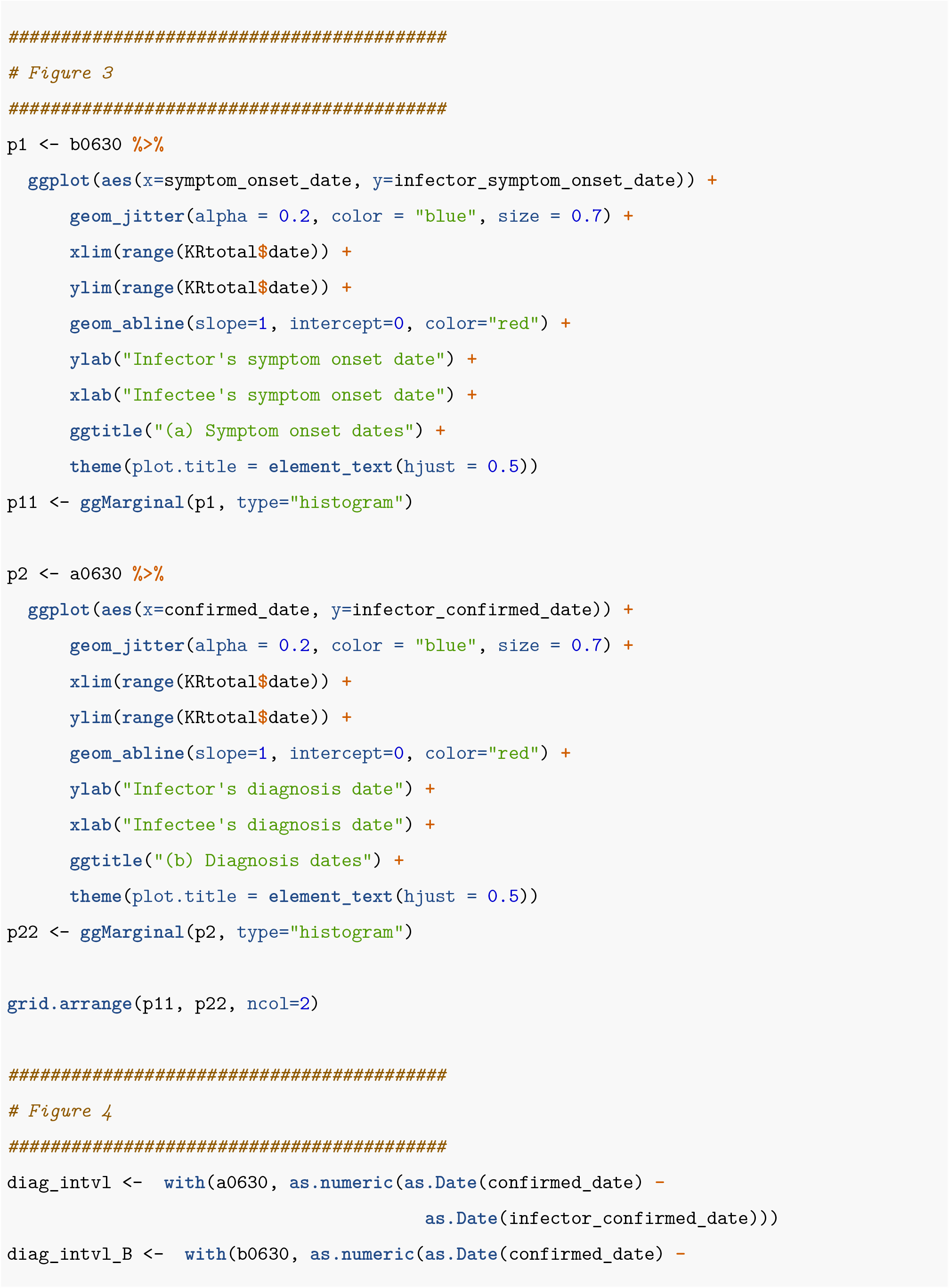
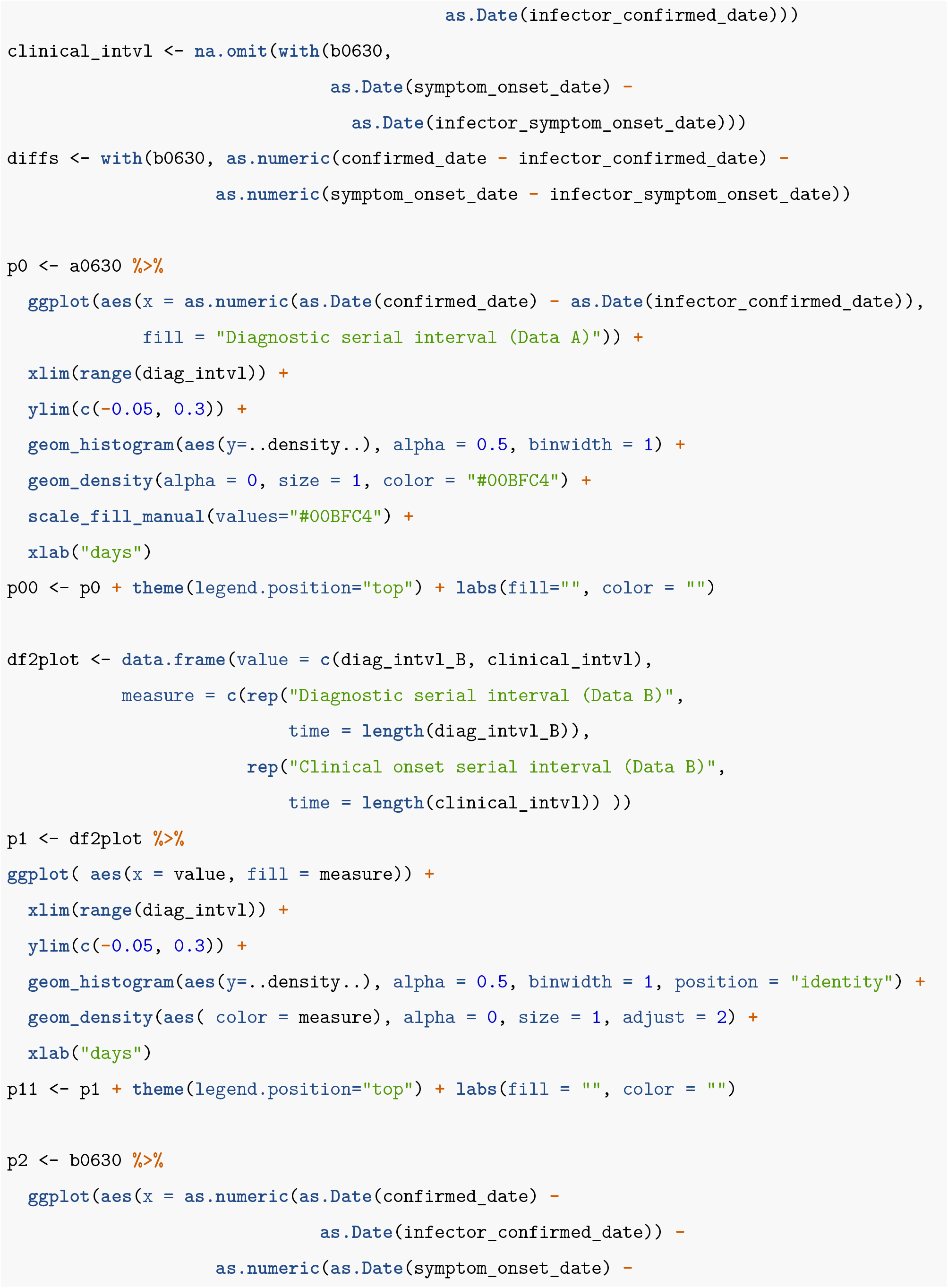
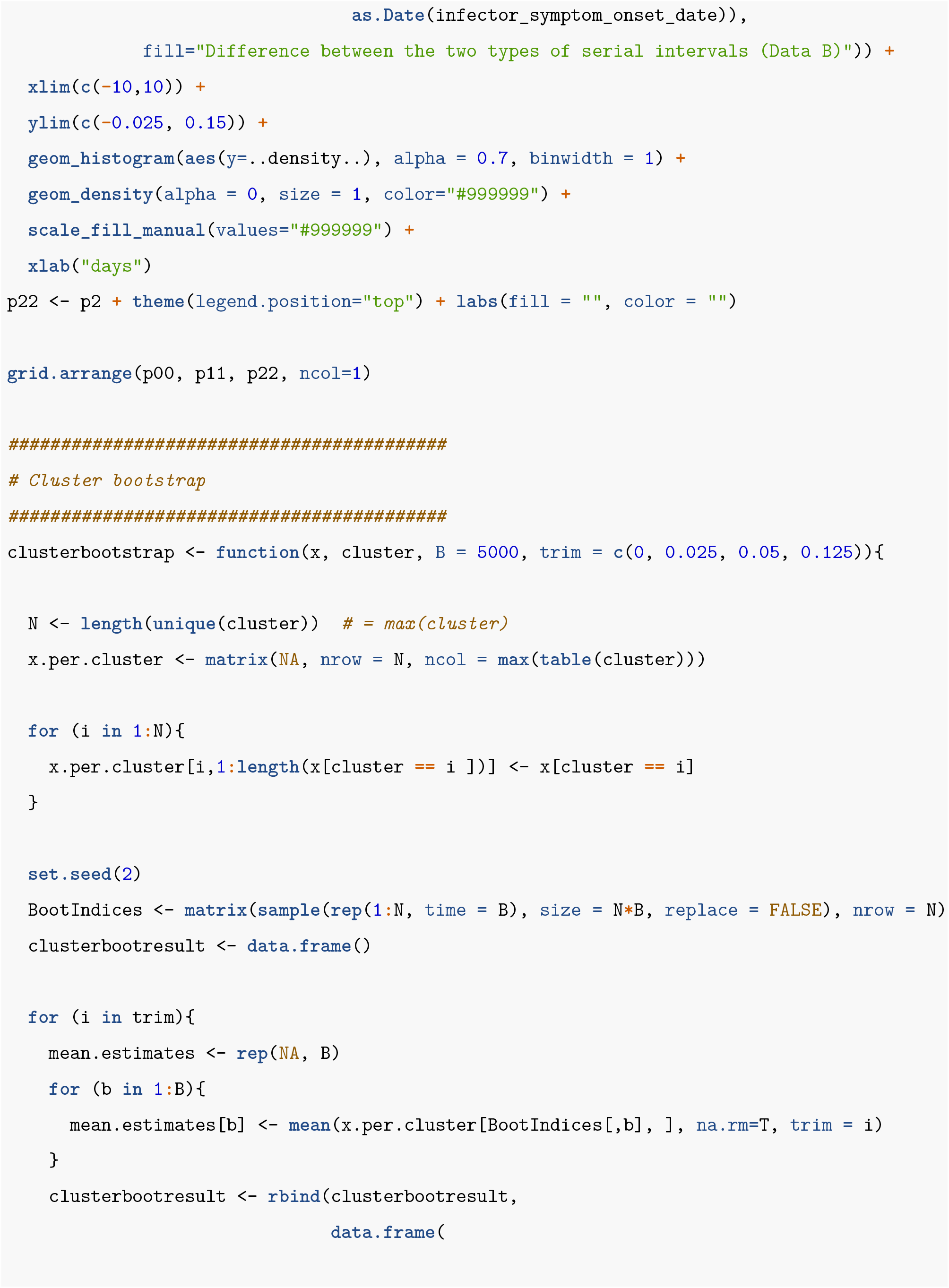
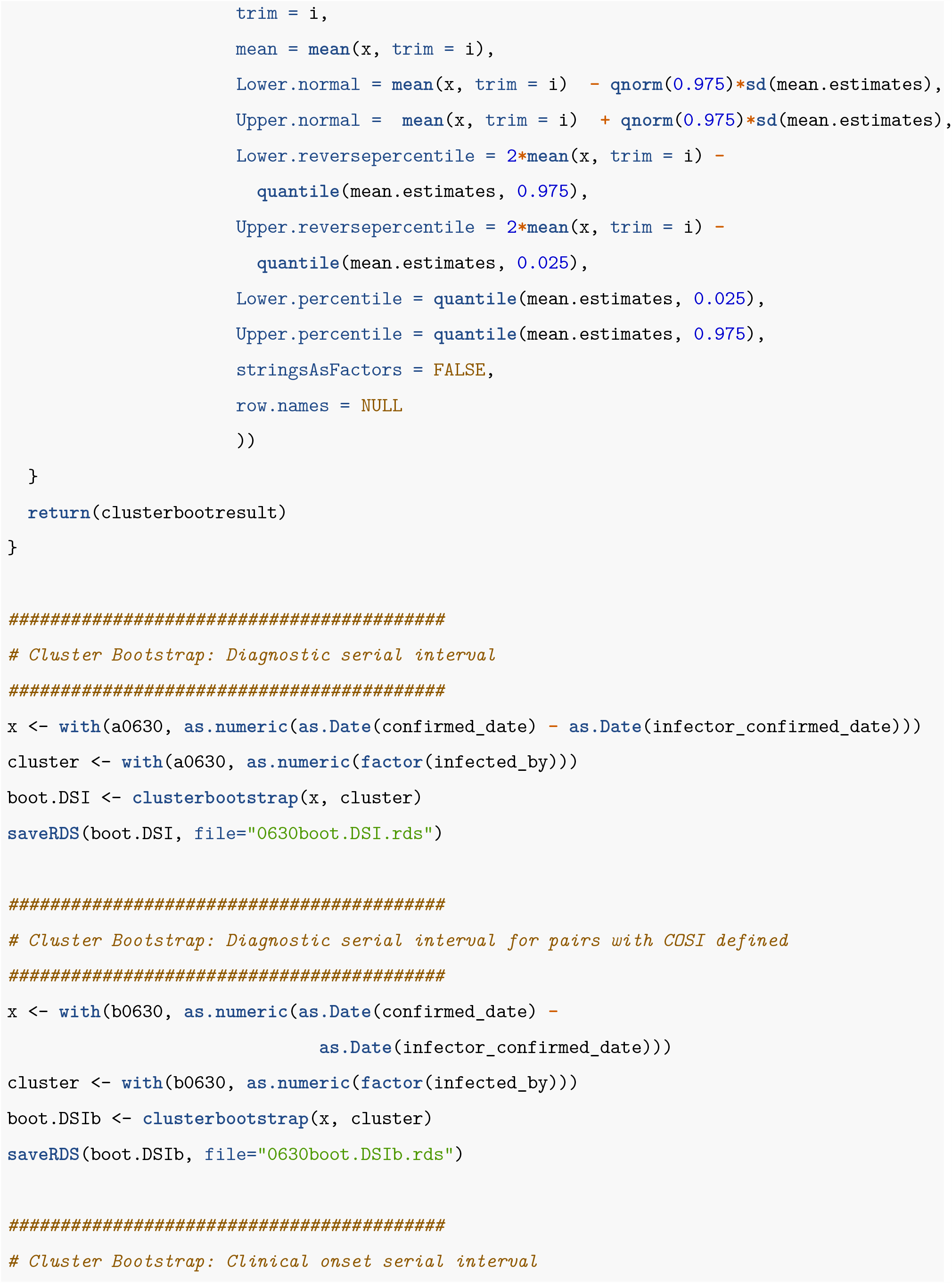
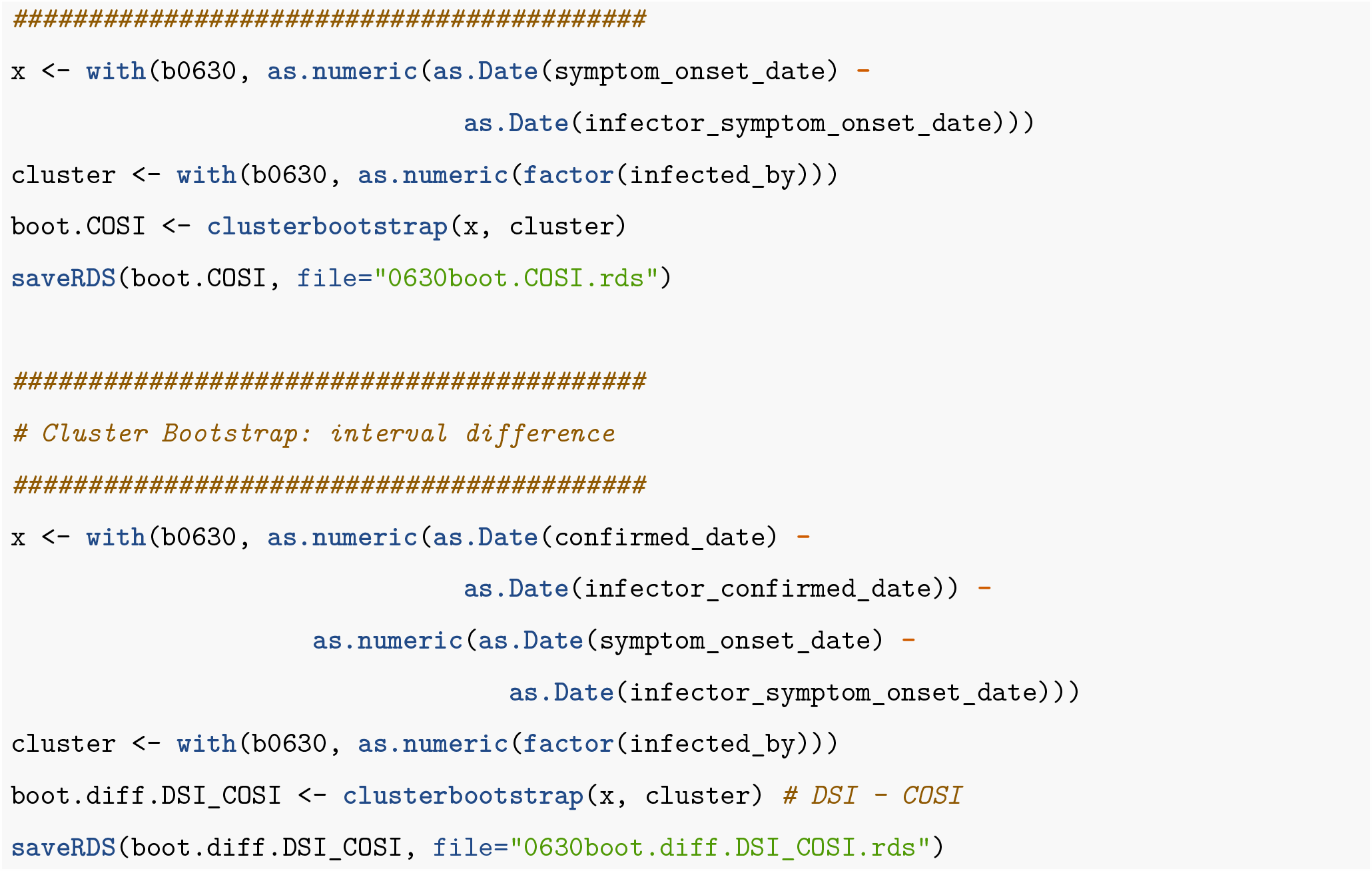

## Notes

### Competing Interest Statement

The authors have declared no competing interest.

### Funding Statement

We declare no funding sources for writing this manuscript.

